# Decrease of implantable cardioverter-defibrillator shock therapy in children: correlation with ICD programming and remote monitoring

**DOI:** 10.1101/2025.04.08.25325494

**Authors:** Robin A. Bertels, Eric Boersma, Martijn D. Zeggelaar, Sophie A.F. van Dongen, Arend D.J. ten Harkel, Lieselot van Erven, Cynthia Smeding, Beatrijs Bartelds, Gert van den Berg, Ewout P. Boesaard, Rohit E. Bhagwandien, Sing-Chien Yap, Reinoud E. Knops, Nico A. Blom, Janneke A.E. Kammeraad

## Abstract

**Background:** Implantable cardioverter defibrillator (ICD) therapy is effective in preventing sudden cardiac death in children. Unnecessary shocks should be avoided. ICD programming strategies and remote monitoring effective in preventing ICD shock therapy in adults have been applied in children

**Objectives:** To investigate the effect of ICD programming and remote monitoring on th incidence of ICD shock therapy in children.

**Methods:** Retrospective multi-center study, including children with transvenous or epicardial ICD implantation. During follow-up ICD-shocks, programming variables and use of remote monitoring were collected.

**Results:** One-hundred-sixteen children were included, median age 13.4 years (min-max 0.3-18), median follow-up 5.2 years (IQR 3.7-6.6). Fifty-three with an ICD implanted *before* 2010 and 63 *after* 2010. The total, appropriate and *in*appropriate annual shock rate decreased from 10.5% to 8% (difference in mean cumulative function (MCF) P=0.008), 7.8% to 5.8% (MCF P=0.036) and 4.3% to 2.6% (MCF P=0.28) respectively, without increase in cardiac related death. The VF zone was programmed higher (≥210 bpm) in patients *before* 2010 compared to *after* 2010 (76% vs 90%; P=0.142), in patients with versus without shocks (79% vs 89%; P=0.243) and at time of appropriate versus *in*appropriate shocks (86% vs 79%; P=0.0016). Remote monitoring was associated with a decrease of total shocks (MCF P=0.013) and appropriate shocks (MCF P=0.052).

**Conclusions:** The incidence of ICD shocks has significantly decreased in children with implantation *after* 2010. A higher programmed VF-zone and application of remote monitoring are correlated to this decrease and therefore justify its use in strategies to prevent unnecessary shocks in children.

## Introduction

Implantable cardioverter defibrillator (ICD) therapy is effective in preventing sudden cardiac death caused by life threatening arrhythmia in children. The ICD indications and programming strategies for children are extrapolated from adult trials,^1,2^ there are no randomized trials conducted in children.^3^ Children requiring ICD therapy for primary and secondary prevention have a variety of underlying diseases, including hypertrophic and dilated cardiomyopathy, primary electrical disease and/or aborted sudden cardiac death, and a broad spectrum of congenital heart disease.^3–5^

Several single center and multi-center retrospective ICD studies and prospective ICD registries in children report appropriate shock rates ranging from 19 to 31% (depending on follow-up duration) and almost no deaths attributable to arrhythmia or device failure.^4,6–10^ However, the complication rate is still significant: *in*appropriate ICD shocks, infections, lead related problems and ICD failure are most common.^11–15^ The incidence of *in*appropriate shocks in children ranges from 20 to 27% during a mean follow up of 3.3 to 5.3 years,^4,6,9,10^ and has negative effect on the psycho-social wellbeing of young patients and their parents.^16,17^ More recent literature tends to report a decrease of *in*appropriate shock rate to around 10%.^11,13,14,18,19^ The incidence of ICD shock therapy over time has not been investigated in children.

It is of utmost importance to minimize unnecessary ICD shock therapy. Technical advances in ICD lead characteristics, advancements in programming strategies of the ICD^20–22^ and the introduction of remote monitoring^23–25^ have led to a reduction in the number of *in*appropriate shocks in adults. These principles are also introduced in the pediatric population. The advantage of early recognition of lead dysfunction and supraventricular or sinustachycardia by remote monitoring is expected to be beneficial particularly for the actively moving pediatric population for the prevention of inappropriate ICD shocks. However, only few studies have assessed the effect of ICD programming^11,13^ and remote monitoring on the ICD shock rate in children.^26–30^ As a consequence specific guidelines describing pediatric ICD programming are not available, but could be useful, as guideline concordant programming in adults has led to a further reduction of ICD shock therapy and mortality.^31,32^

The current study investigates the incidence of ICD shock therapy in a Dutch cohort of pediatric ICD patients over time and the effect of ICD programming strategies and remote monitoring on the incidence of appropriate and *in*appropriate ICD shocks in children.

## Methods

This retrospective multi-center study included all pediatric patients (0-17 years of age), who had a transvenous or epicardial ICD implantation between 1995 and 2021 in one of three University Medical Centers in the Netherlands, regardless of the underlying diagnosis. Patients with a subcutaneous ICD system were excluded. An official waiver of ethical approval was granted from the local ethical committees.

### Baseline characteristics

Patient baseline characteristics at ICD implantation were obtained from digital patients files: year of implant, age, sex, weight, diagnosis, primary or secondary prevention and implantation route. ICD programming variables were collected directly after implantation, at one month follow up and at most recent follow up. VF/fast-VT/VT zones and detection time were categorized to be able to perform statistical analysis; VF zone categorized in < 210/min, 210-230/min, ≥ 230/min, (fast-)VT zone categorized in < 190/min, 190-210/min, ≥ 210/min, detection times categorized < 3 sec, 3-6 sec, ≥ 6 sec. For comparison purposes, the detection time recorded as number of beats were converted to time in seconds. Supraventricular tachycardia (SVT) discriminators, programmed anti-tachycardia pacing (ATP) therapy and starting date of remote monitoring (at the implantation or during follow-up) were registered.

### Follow-up

During follow-up, all ICD shock episodes were collected, together with the ICD programming variables at the time of and after a shock episode. An ICD shock episode was defined as one or more ICD shocks until redetection of sinus rhythm by the device. To improve readability, one ICD therapy episode will be referred to as one ICD shock. *In*appropriate shocks were defined as ICD therapy for other tachycardia (SVT or sinus tachycardia) or for an abnormal sensing episode. Adverse events such as the need for device or lead revision and sudden cardiac arrest (SCA) requiring cardiopulmonary resuscitation (CPR), were registered separately. Follow-up continued until patients were 18 years of age or had at least a 5-year follow-up after they reached 18 years of age.

### Endpoints

The primary end-points were differences in (F)VT/VF zones, detection time, SVT discriminator programming and the use of remote monitoring in 1) patients with an ICD implantation up to and including 2010 (*before* 2010) versus *after* 2010; 2) patients with ICD shocks versus without ICD shocks; 3) in relation to the proportion of appropriate versus *in*appropriate shocks. The cut-off year 2010 was chosen to divide the patient cohort in two groups, as it marks the period around when data on the Dutch pediatric ICD patient cohort were published,^4^ and when adult ICD studies concerning specific programming strategies led to changes in ICD management for children. ^21,25^ After 2010 remote monitoring got widely implemented.

### Statistical analysis

Continuous data are presented as mean ± standard deviation or median (interquartile range), depending on the normality of the distribution, which was evaluated by the Shapiro-Wilk test. Differences between the *before* and *after* 2010 cohorts were analyzed using an independent t-test (for data with normal distribution) or Mann-Whitney U test. Categorical data are presented as an absolute count and percentage, and differences between the cohorts were evaluated by Chi-square tests or Fisher’s exact tests (in case of an expected value <5).

The number of shocks, appropriate shocks and *in*appropriate shocks are presented as a (non-parametric) mean cumulative function (MCF) as this method facilitates multiple event (shocks) modelling.^33^ Differences in MCF between the *before* and *after* 2010 cohorts, and between patients with versus without remote monitoring (at any time prior to the shock and the two cohorts taken together) were analyzed by a chi-square test according to the method of Nelson.^34^ Differences between the cohorts in the time to the first occurrence of a shock (total, appropriate, *in*appropriate) were analyzed using the Cox proportional hazard regression model. Univariable models were ran, and models with multivariable adjustment for age and sex, and additionally for remote monitoring (as a time-dependent covariate), primary versus secondary prevention, and transvenous versus epicardial implantation. Results are presented as hazard ratios (HRs) with corresponding 95% confidence interval (CI). Statistical analysis was performed using SAS Analytics Software version 9.4. A p-value of < 0.05 was considered significant.

## Results

A total of 163 children received a transvenous or epicardial ICD implantation, 88 patients *before* 2010 and 75 patients *after* 2010. Sufficient retrospective data could be retrieved of 116 patients to be able to include them in this study, 53 patients *before* 2010 and 63 patients *after* 2010. The baseline characteristics are presented in table 1. The median age at implantation was 13.4 years of age, with no difference in age *before* and *after* 2010. The male to female ratio was 62 – 38 %. The ICD indications did not change over the years, with an almost 50-50 distribution of primary versus secondary prevention.

**Table 1:** Baseline characteristics.

|  | ICD implantation<br>*before 2010* | ICD implantation<br>*after 2010* | Total | P-value |
| --- | --- | --- | --- | --- |
| Number of patients, # | 53 | 63 | 116 |  |
| Sex (male), # (%) | 35 (66) | 37 (59) | 72 (62) | 0.419 |
| Age (years), median (min-max) | 14.2 (0.3-18.0) | 13.1 (0.3-17.7) | 13.4 (0.3-18) | 0.595 |
| Weight (kg), mean (SD) | 46.5 (+/- 23.7) | 44.0 (+/- 22.4) | 45.0 (+/- 22.9) | 0.870 |
| Diagnosis, # (%)<sup>a</sup> |  |  |  |  |
| - HCM | 15 (28) | 12 (19) | 27 (23) |  |
| - CPVT | 6 (11) | 3 (5) | 9 (8) |  |
| - LQT | 4 (8) | 8 (13) | 12 (10) |  |
| - VF/VT idiopathic | 11 (21) | 13 (21) | 24 (21) |  |
| - DCM | 8 (15) | 11 (18) | 19 (16) |  |
| - Other<sup>‡</sup> | 9 (17) | 16 (25) | 25 (22) |  |
| Prevention, # (%) |  |  |  |  |
| - primary | 26 (49) | 33 (52) | 59 (51) | 0.721 |
| - secondary | 27 (51) | 30 (48) | 57 (49) |  |
| Route, # (%) |  |  |  |  |
| - transvenous | 44 (83) | 47 (75) | 91 (78) | 0.272 |
| - epicardial | 9 (17) | 16 (25) | 25 (22) |  |
| Pacing mode, # (%) |  |  |  |  |
| - DDD | 15 (28) | 26 (41) | 41 (35) | 0.069 |
| - VVI | 33 (62) | 27 (43) | 60 (52) |  |

CPVT = catecholaminergic polymorphic ventricular tachycardia; DCM = dilated cardiomyopathy; HCM = hypertrophic cardiomyopathy; LQT = long QT syndrome; VF/VT = ventricular fibrillation / tachycardia.
<sup>a</sup> A patient can have a diagnosis in more than one category, but is counted in only one category
<sup>b</sup> Other: arrhythmogenic right ventricular cardiomyopathy, restrictive cardiomyopathy, rhabdomyoma, non- compaction cardiomyopathy, myocardial infarction, Brugada syndrome.

### Incidence of ICD shock therapy

Fifty-five out of 116 patients (47%) received a total of 216 shocks, with a median of 2 shocks per patient (range 1–29 shocks) during a median follow-up of 5.2 years (IQR 3.7–6.6 years, Table 2). The number of patients with an ICD shock decreased from 30 (57%) *before* 2010 to 25 (40%) *after* 2010. The difference in MCF of ICD shocks per patient was significant (P = 0.008, Figure 1). With a mean follow-up of 5.4 years and 5.0 years respectively, the annual ICD shock rate decreased from 10.5% *before* to 8.0% *after* 2010.

**Table 2:** Number of patients with or without shocks and types of shock.

|  | ICD implantation<br>*before 2010* | ICD implantation<br>*after 2010* | Total | P-value |
| --- | --- | --- | --- | --- |
| Number of patients, # | 53 | 63 | 116 |  |
| Follow-up time (years),<br>median (IQR) | 5.4 (4.8 - 7.0) | 5.0 (2.8 - 5.9) | 5.2 (3.7 - 6.6) | 0.007 |
| Patients with shocks, # (%) | 30 (57) | 25 (40) | 55 (47) | 0.069 |
| - appropriate shocks, # (%) | 22 (42) | 18 (29) | 40 (35) | <sup>a</sup> |
| - inappropriate shocks, # (%) | 12 (23) | 8 (13) | 20 (17) | <sup>a</sup> |
| - unknown shocks, # (%)<sup>b</sup> | 8 (15) | 0 (0) | 8 (7) | <sup>a</sup> |

<sup>a</sup> A patient can have had shocks in more than one category, therefore statistical tests were performed in the mean cumulative number of shock curves (Figure 1).
<sup>b</sup> Shocks with insufficient data to define the appropriate or inappropriate origin of the shock.

**Figure 1.**
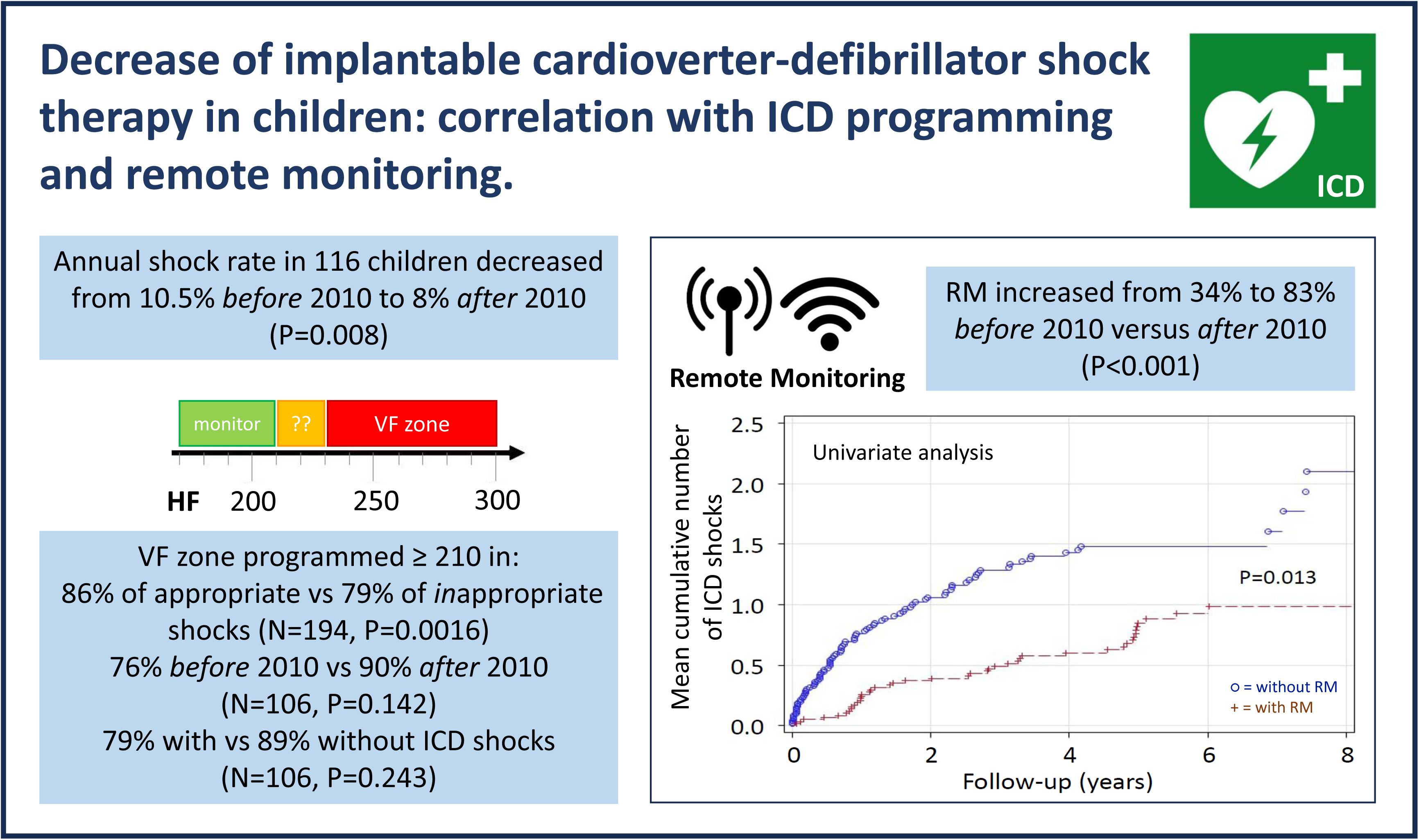
Mean cumulative number of shocks per patient with ICD implant up to and including 2010 (blue line) or *after* 2010 (red dotted line)

A similar significant decrease was observed in the MCF of appropriate shocks per patient (P = 0.036, Figure 1). The number of patients with appropriate shocks decreased from 22 (42%) to 18 (29%), and the annual appropriate shock rate from 7.8% *before* 2010 to 5.8% *after* 2010. The proportion of patients that received an *in*appropriate shock decreased from 23% *before* to 13% *after* 2010, the annual *in*appropriate shock rate decreased from 4.3% to 2.6%. The difference in MCF of *in*appropriate shocks per patient was not significant (P = 0.28 Figure 1).

### ICD programming

Comparing the VF detection zone programming at the time of shock between appropriate and *in*appropriate shocks showed significant higher VF zone programming (≥ 210/min) in appropriate shocks (86% vs 79%; P = 0.0016, Table 3). Comparing the programmed VF detection zones in patients implanted before and after 2010 did not show a significant difference, but a trend towards higher programmed VF zones (≥ 210 BPM) at end of study follow up in 90% of patients with ICD implantation *after* 2010 versus 76% of patients implanted *before* 2010 (P = 0.142 Table 1 Supplementary material). Comparing patients with and without shocks also showed a trend towards higher programmed VF zones (≥ 210 BPM) at end of study follow up in the patients without shocks (79% vs 89%; P=0.243, table 3). The detection time programming in the VF zone was not different in the group with versus without shocks and in appropriate versus *in*appropriate shocks (Table 3). The (fast-)VT zone was used for ICD therapy in 34% (39/116) of patients, 38% *before* and 30% *after* 2010 (Table 1 Supplementary material), with no change in detection rate *before* versus *after* 2010.

**Table 3:**
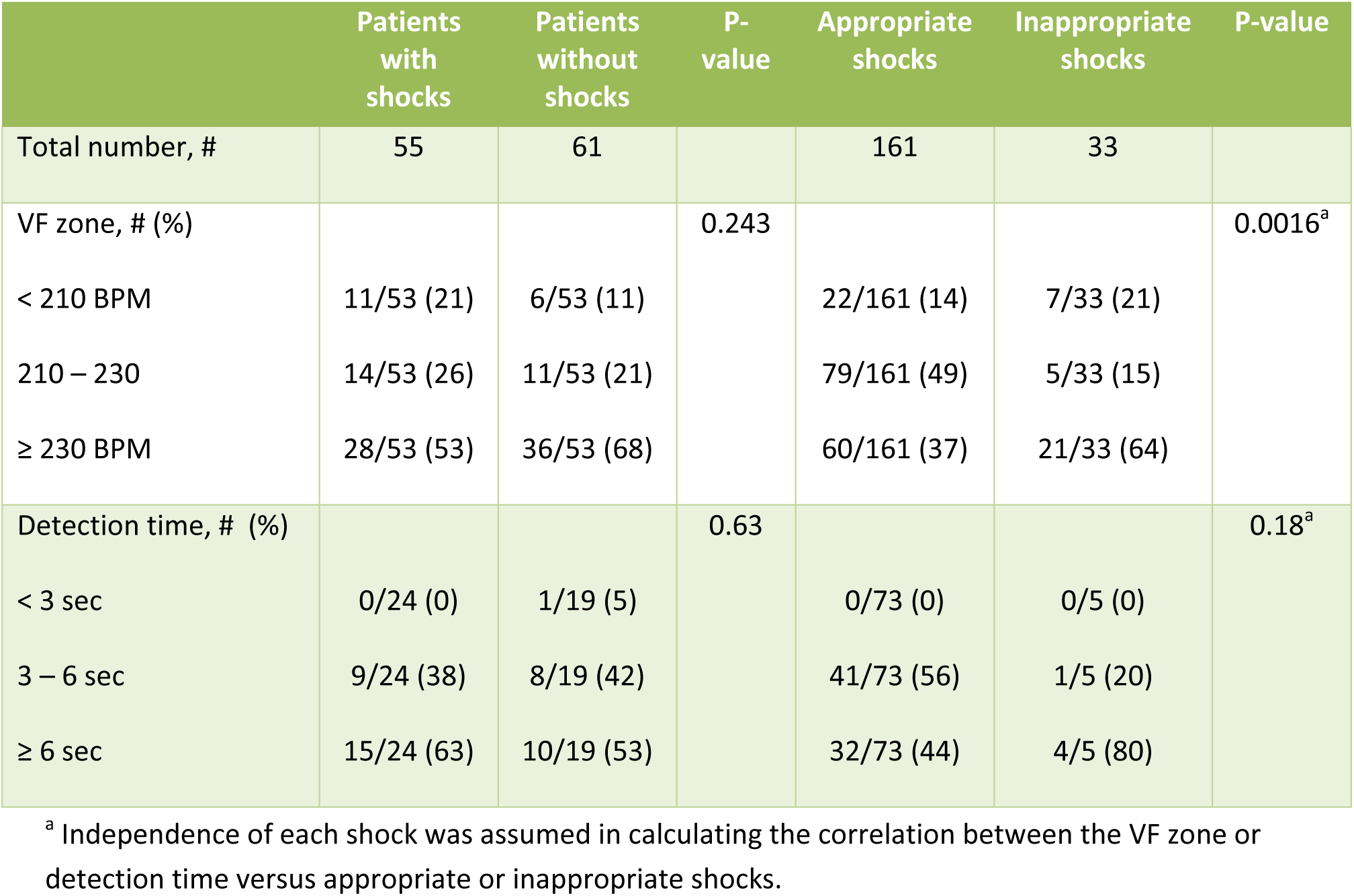
ICD programming in patients with shocks versus without shocks at end of study follow-up; and at time of appropriate shocks versus *in*appropriate shocks.

|  | Patients with shocks | Patients without shocks | P-value | Appropriate shocks | Inappropriate shocks | P-value |
| --- | --- | --- | --- | --- | --- | --- |
| Total number, # | 55 | 61 |  | 161 | 33 |  |
| VF zone, # (%) |  |  | 0.243 |  |  | 0.0016<sup>a</sup> |
| < 210 BPM | 11/53 (21) | 6/53 (11) |  | 22/161 (14) | 7/33 (21) |  |
| 210 – 230 | 14/53 (26) | 11/53 (21) |  | 79/161 (49) | 5/33 (15) |  |
| ≥ 230 BPM | 28/53 (53) | 36/53 (68) |  | 60/161 (37) | 21/33 (64) |  |
| Detection time, # (%) |  |  | 0.63 |  |  | 0.18<sup>a</sup> |
| < 3 sec | 0/24 (0) | 1/19 (5) |  | 0/73 (0) | 0/5 (0) |  |
| 3 – 6 sec | 9/24 (38) | 8/19 (42) |  | 41/73 (56) | 1/5 (20) |  |
| ≥ 6 sec | 15/24 (63) | 10/19 (53) |  | 32/73 (44) | 4/5 (80) |  |

<sup>a</sup> Independence of each shock was assumed in calculating the correlation between the VF zone or detection time versus appropriate or inappropriate shocks.

### Remote monitoring

Remote monitoring was applied during follow-up in 18/53 patients (34%) *before* 2010 and increased to 52/63 patients (83%) *after* 2010 (P < 0.001). The MCF of total shocks in patients with and without remote monitoring showed a hazard ratio of 0.59 (95% CI 0.41 – 0.85, P = 0.013, Figure 2) in patients with remote monitoring applied. For the MCF of appropriate ICD shocks the hazard ratio was 0.61 (95% CI 0.40 – 0.94) in patients with remote monitoring applied (P = 0.052). For *in*appropriate shocks the MCF did not show a significant difference with and without remote monitoring.

**Figure 2.**
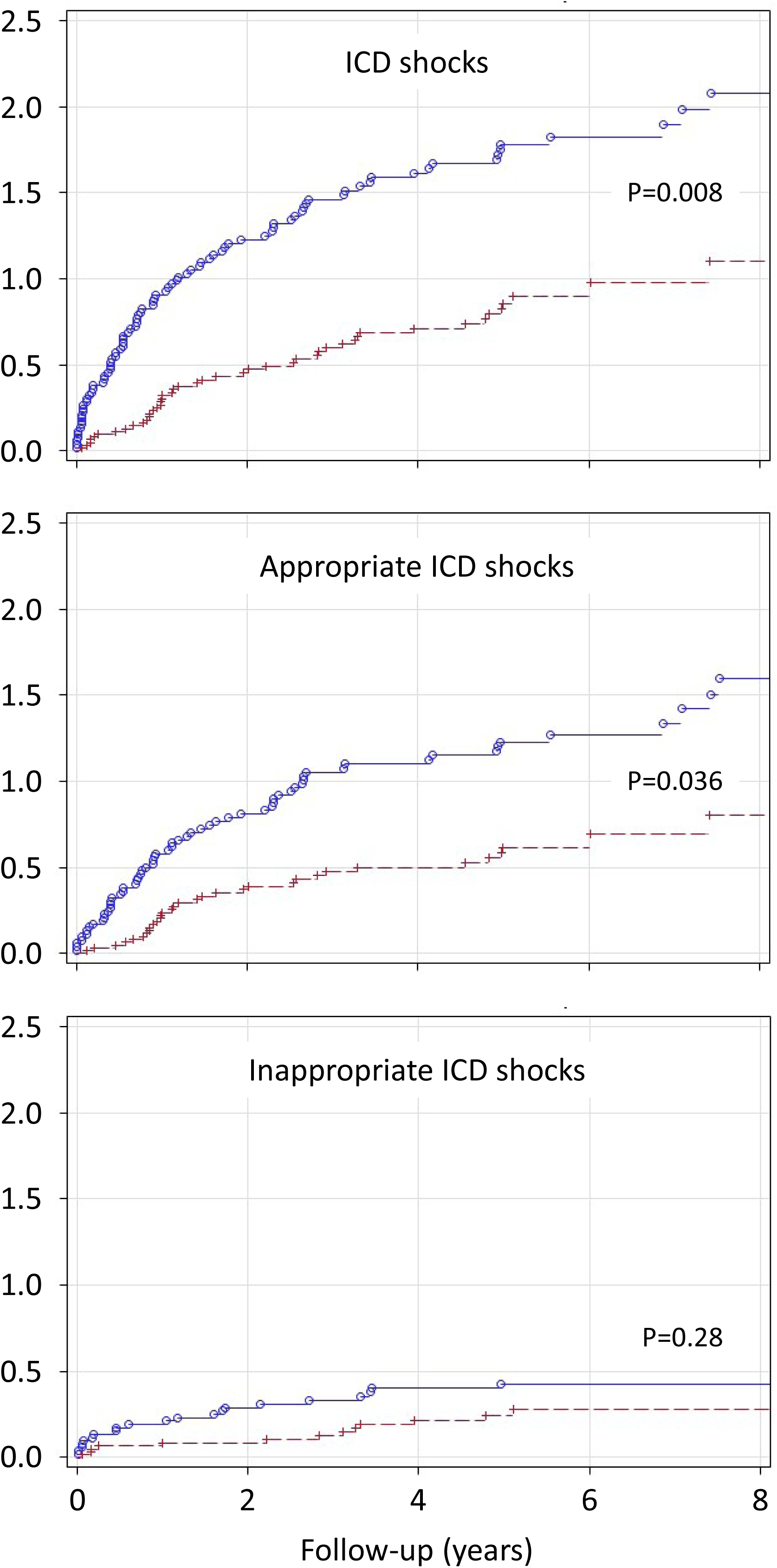
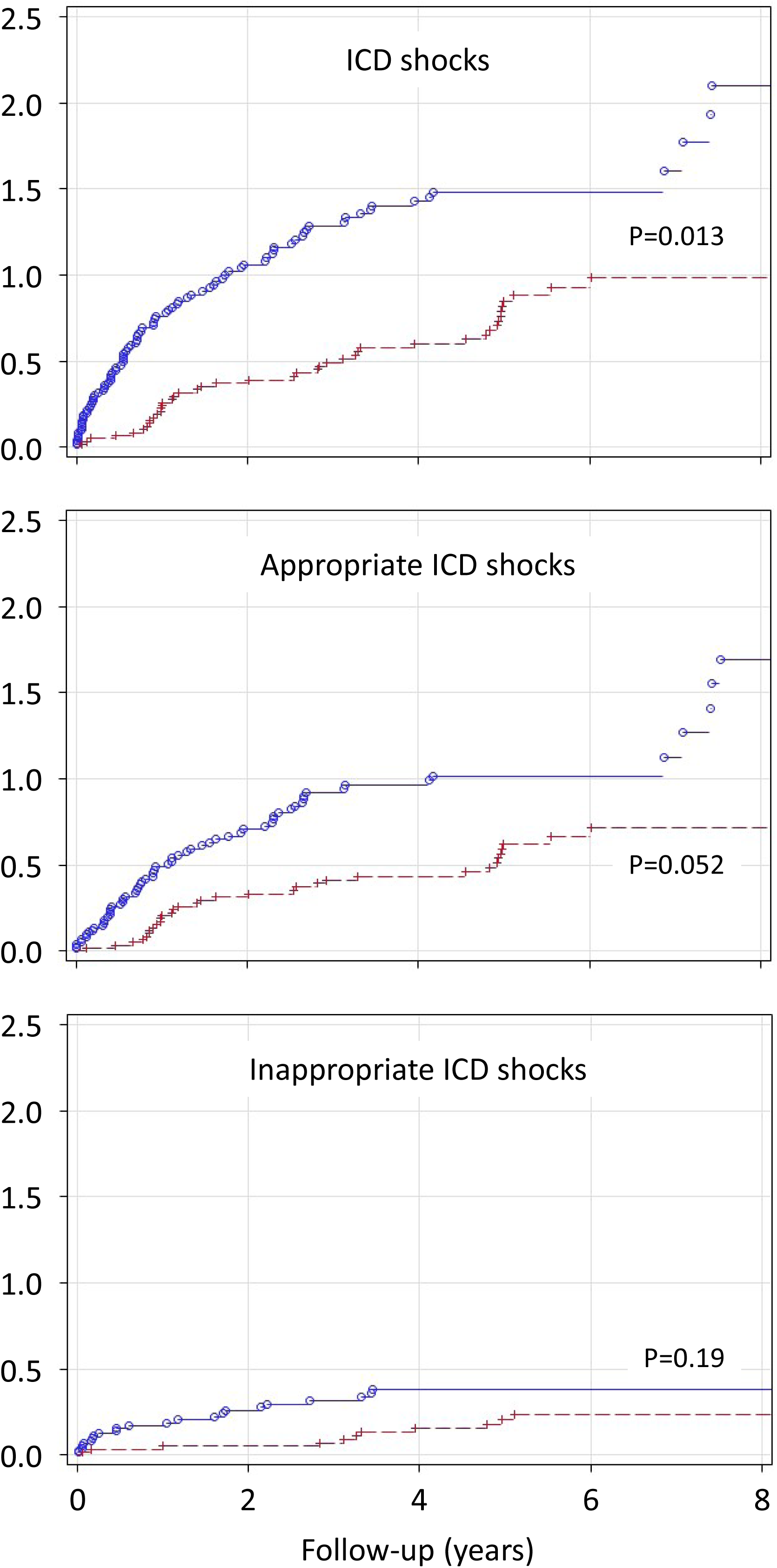
Mean cumulative number of shocks per patient with remote monitoring applied (red dotted line) or with remote monitoring not applied (blue line)

In explorative multivariate analysis, ‘implantation *after* 2010’ and ‘older age’ appeared to be negatively correlated with total (Table 4) and appropriate ICD shocks. Surprisingly, in multivariate analysis of *in*appropriate shocks secondary prevention was positively correlated with *in*appropriate ICD shocks, hazard ratio 2.96 (95% CI 1.39 – 6.32).

**Table 4:** Hazard ratio in multivariate analysis of total ICD shocks.

|  | Hazard ratio | 95% confidence interval |
| --- | --- | --- |
| Implantation date (*after* 2010) | 0.429 | 0.240 – 0.767 |
| Age (years) | 0.920 | 0.874 – 0.969 |
| Gender (male) | 1.009 | 0.698 – 1.459 |
| Remote monitoring (applied) | 1.160 | 0.649 – 2.072 |
| Prevention (secondary) | 1.398 | 0.980 – 1.993 |
| Route (transvenous) | 0.992 | 0.535 – 1.839 |

### Complications of ICD treatment

During a mean follow-up of 5.2 years, 39 patients (34%) underwent an ICD or lead revision. Reasons for ICD revision were end-of-life of the battery in 19 patients, lead dysfunction in 17 patients, and infection and other indications in 8 patients. Some patients had a combination of reasons for revision.

Three patients had an aborted sudden cardiac arrest requiring CPR despite ICD therapy, all with implantation *after* 2010. Two patients had a VT under the detection limit of 222/min (VF zone) and 231/min (fast-VT zone, VF-zone > 250/min) respectively, which was not hemodynamically tolerated; in a third patient with a VF zone > 240/min VF was detected, but only the fourth shock was effective.

Twelve patients underwent a heart transplantation during follow-up, 5 patients *before* 2010 and 7 patients *after* 2010. In the group of patients *before* 2010, three patients died during follow-up of the study, compared to one patient in the group *after* 2010. The cause of death was cardiac, but unrelated to ICD therapy in 3 patients and unknown in another patient.

## Discussion

### Main findings

This study demonstrates a significant decrease in total amount of ICD shocks over the last decade and a protective correlation of remote monitoring on the incidence of ICD shocks in one of the largest cohorts of pediatric patients. Contrary to expectations, this decrease and protective effect was clearly present for appropriate shocks, but could not be demonstrated for *in*appropriate shocks. In addition, the VF detection rate was significantly more often programmed in higher ranges (≥210/min) at time of appropriate shocks when compared to *in*appropriate shocks. Also a trend towards higher VF detection zone (≥210/min) *after* 2010 and in patients without shocks was demonstrated.

### Annual shock rate

The annual appropriate shock rate in this study was 7.8% and 5.8%, *before* 2010 and *after* 2010 respectively. The annual appropriate shock rate *before* 2010, matches the previously reported annual appropriate shock rate of 8.4% in the Netherlands, which included partly the same patients.^4^ The annual appropriate shock rate *after* 2010 is in range with other pediatric ICD series published over the last 10 years (Table 2 Supplementary Material), describing annual appropriate shock rates ranging between 2.9 and 13.9%. Also the annual *in*appropriate shock rate of 2.6% *after* 2010 is in line with the annual *in*appropriate shock rate in the review varying between 1.1 to 6.8% in recent studies. These recently described appropriate and *in*appropriate shock rates in children approximate the numbers described for adults with inherited arrhythmia syndromes. In a meta-analysis of patients with a mean age of 39 years and inherited arrhythmia syndromes, the annual rate of *in*appropriate shocks was 4.7%.^35^

### Influence of ICD programming

This study describes a decrease of ICD shocks over the last decade, with a trend towards higher detection rates (≥210 bpm) in VF zones *after* 2010. In addition, this study shows a significant higher programmed VF zone in appropriate shocks in comparison to *in*appropriate shocks. Both findings confirm the advantage of high VF zone programming. In a study from 2015 including pediatric and adult congenital heart disease patients with an ICD, with a comparable annual appropriate ICD shock rate of 5.7%, had a mean ventricular fibrillation detection rate of 222 +/− 15 beats/min.^8^ It showed no difference in patients with or without *in*appropriate shocks, by comparing ICD programming data at the time of *in*appropriate shock to programming data of the patients without *in*appropriate shock at most recent follow up. A more recent study in 51 pediatric patients from 2023 did show a difference in programmed VF zone in patients with *in*appropriate shocks (VF zone 188/min) vs patients without *in*appropriate shocks (VF zone 222/min).^13^ The inconsistency of these findings might partly be attributed to the potential incomparability of data measured at the time of the *in*appropriate shock and at random follow up time in patients without *in*appropriate shocks. This was the reason not to perform these analyses in the current study. However, the results of previous studies, as well as the results described in this study, justify the current practice to program VF detection rates up to 230-250/min in children, in accordance to adult guideline on ICD programming.^32^ These recommendations in adults are, amongst other studies, based on large trials like the MADIT-RIT and RISSY-ICD trials, which have clearly shown the beneficial effects of high-rate programming with VF-zones higher than 200-230/min on *in*appropriate shocks and mortality.^20,21^

A longer detection time is also known to decrease the shock rate in adult patients.^22^ In the ADVANCE III trial in adult patients the appropriate and *in*appropriate shock rate decreased 48% by increasing the detection time from 18 out of 24 intervals, to 30 out of 40 intervals.^36^ Though this effect has not been confirmed in pediatric series, some studies do report the use of longer detection times, like a median detection time of 9.6 seconds in the UK National hypertrophic cardiomyopathy (HCM) cohort study.^11^ The current study was unable to demonstrate an effect of the detection time on the total, as well as the appropriate or *in*appropriate shock rate.

Monomorphic ventricular tachycardia has been found to be a risk factor for appropriate shocks in pediatric patients.^13^ Therefore, therapy in VT-zone (ATP and shock) is often programmed in pediatric patients with prior documented VT episodes.^11^ On the other hand, studies in adults and children have shown that the use of shock therapy in the VT-zone increases appropriate and *in*appropriate shocks, and even increases all-cause mortality.^8,21^ In our present series we found that therapy in the (fast-)VT-zone was programmed in 34% of the patients, without a difference in detection rate between the patients with implantation *before* or *after* 2010. As the current study did not collect data on whether shock therapy was applied in the (fast-)VT or VF zone, the relation between (fast-)VT zone programming and ICD shock rate could not examined.

### Influence of remote monitoring

Remote monitoring was significantly associated with a reduction in total amount of ICD shocks. Although remote monitoring was expected to be particularly advantageous for the prevention of *in*appropriate ICD shocks, its efficacy was only confirmed for a decrease in any shock and appropriate shocks. In the adult literature, there is clear evidence that both any ICD shock and *in*appropriate shocks can be reduced by 72% and 52% respectively with remote monitoring.^23^ This finding was confirmed by a meta-analysis in 2015.^37^ The evidence in literature for the additional value of remote monitoring in the prevention of ICD shocks in pediatric patients with an ICD is scarce.^38^ A study from 2014 with 198 patients with a cardiac implantable electronic device (CIED) of which 61 ICD, showed that remote monitoring allows for early identification of arrhythmias (like SVT, atrial fibrillation/flutter or VT) and device malfunction.^28^ But there is no previously published evidence that remote monitoring correlates with a reduction of the ICD shock rate in a pediatric population (Table 2 Supplementary Material).

In univariate analysis remote monitoring was associated with ICD shock reduction over the last decade. Multivariate analysis showed that only ‘implantation *after* 2010’ and ‘older age’ were negatively correlated with the amount of total and appropriate ICD shocks. This suggests that other factors also must have contributed to ICD shock reduction. A combination of technical advances in ICD lead preservation and ICD programming management probably played an important role, as for example the trend to higher detection rate programming described in this study. In addition, advances and new insights to disease treatment will have contributed to a decrease of the incidence of arrhythmia or ICD implantation in general.^39,40^ These factors were not investigated in this study.

### Other risk factors for shocks

In the current study a younger age was correlated with a higher proportion of total shocks and appropriate shocks in multivariate analysis. We did not find a correlation between age and *in*appropriate shocks. Evidence on the effect of age on the number of shocks in the literature is conflicting (Table 2 Supplementary Material), some do report a correlation with appropriate shocks and younger age,^4^ while others do not find any association.^10^ The same accounts for the correlation between *in*appropriate shocks and age.^7,8^ In general, an ICD indication at younger age is reserved for the most serious disease phenotypes, with consequently the highest arrhythmia burden and higher shock rates.

Secondary prevention has been identified as a predictor of appropriate shocks in adult patients,^41^ but was described with inconsistent results in pediatric series (Table 2 Supplementary Material). Unexpectedly, multifactorial analysis in the current series, showed a correlation between secondary prevention and increased risk for *in*appropriate shocks. Since this has not been reported in other series, this result should be interpreted with caution.

### Implications for clinical practice

Based on the results of the present study and the review of the literature, we conclude that higher VF detection zone programming and the application of remote monitoring are beneficial for children, and can reduce the number of appropriate and *in*appropriate shocks. The current study supports the concept to program pediatric ICDs with high VF zones above 210-230/min, with the application of remote monitoring. Exceptions to standard high rate VF/(fast-)VT zone programming are patients with ventricular dysfunction and/or patients who do not tolerate slow VT hemodynamically. In the current study ICD programming data were not related to ventricular ejection fraction or diastolic dysfunction. However, two patients required CPR despite ICD implantation because of hemodynamically non-tolerated slow VT under the programmed VF/(fast-)VT detection rates. Additional studies are necessary to investigate specific risk factors and ICD programming strategies for pediatric patients with ventricular dysfunction.

### Limitations

This study describes a relatively large cohort of pediatric patients over a long follow-up period. The statistically significant but clinically small difference in follow-up duration *before* and *after* 2010, is not thought to explain the difference in number of shocks *before* and *after* 2010. Due to its retrospective design, available data were often insufficient for reliable data acquisition, particularly in the era < 2010, resulting in inability to investigate all the predetermined end-points (detection time and SVT discriminators). Data collection was focused on the *(in)*appropriateness of ICD shocks, but the exact tachycardia rate and zone for which the specific ICD shock therapy was applied, was not registered. Therefore the influence of (fast-)VT zone programming could not be analyzed, although it might have influenced the shock rates in the study population described. We were also unable to obtain reliable data on the incidence of ATP therapy, anti-arrhythmic drug treatment and left sympathetic cardiac denervation, which could also have affected the shock rate. Finally general conclusions are drawn for a heterogenous group of underlying cardiac diagnosis, but numbers are too low for subgroup analyses. Future large, international (multicenter) prospective studies are needed to overcome the above described limitations.

## Conclusion

The incidence of total amount of ICD shocks and appropriate shocks in children with a transvenous or epicardial ICD, has significantly decreased in patients with implantation *after* 2010 compared to patients with implantation *before* 2010, without an increase of cardiac related death. Factors that are correlated with this decrease in number of shocks are a higher programmed VF-zone and applied remote monitoring. These findings encourage ICD programming of high VF zone detection rates (in children without severe ventricular dysfunction) and the application of remote monitoring in all children requiring ICD therapy.

## Data Availability

The authors confirm that the data supporting the findings of this study are available upon request.

## Abbreviations

SVT: supraventricular tachycardia
VT: ventricular tachycardia
VF: ventricular fibrillation
SCA: sudden cardiac arrest
CPR: requiring cardiopulmonary resuscitation
CIED: cardiac implantable electronic device
ICD: implantable cardioverter defibrillator
ATP: anti-tachycardia pacing
MCF: mean cumulative function

## Acknowledgements

Marco N. Kruit, Arthur E. van der Mark and Michiel Zumbrink for their enormous efforts to provide the availability of data with respect to ICD programming. Mattie J. Lenzen and Danny Ng for technical support with the database. Arend W. van Deutekom for help with preliminary statistical analyses. Reinder Evertz and Leendert van den Beukel for their help to provide long term follow up data.

## References

1. Al-Khatib SM, Stevenson WG, Ackerman MJ, Bryant WJ, Callans DJ, Curtis AB, Deal BJ, Dickfeld T, Field ME, Fonarow GC, et al. 2017 AHA/ACC/HRS Guideline for Management of Patients With Ventricular Arrhythmias and the Prevention of Sudden Cardiac Death: Executive Summary: A Report of the American College of Cardiology/American Heart Association Task Force on Clinical Practice Guidelines and the Heart Rhythm Society. J Am Coll Cardiol. 2018;72:1677–1749. doi: 10.1016/j.jacc.2017.10.053

2. Zeppenfeld K, Tfelt-Hansen J, de Riva M, Winkel BG, Behr ER, Blom NA, Charron P, Corrado D, Dagres N, de Chillou C, et al. 2022 ESC Guidelines for the management of patients with ventricular arrhythmias and the prevention of sudden cardiac death. Eur Heart J. 2022;43:3997–4126. doi: 10.1093/eurheartj/ehac262

3. Writing Committee M, Shah MJ, Silka MJ, Silva JNA, Balaji S, Beach CM, Benjamin MN, Berul CI, Cannon B, Cecchin F, et al. 2021 PACES Expert Consensus Statement on the Indications and Management of Cardiovascular Implantable Electronic Devices in Pediatric Patients: Developed in collaboration with and endorsed by the Heart Rhythm Society (HRS), the American College of Cardiology (ACC), the American Heart Association (AHA), and the Association for European Paediatric and Congenital Cardiology (AEPC) Endorsed by the Asia Pacific Heart Rhythm Society (APHRS), the Indian Heart Rhythm Society (IHRS), and the Latin American Heart Rhythm Society (LAHRS). JACC Clin Electrophysiol. 2021;7:1437–1472. doi: 10.1016/j.jacep.2021.07.009

4. Heersche JH, Blom NA, van de Heuvel F, Blank C, Reimer AG, Clur SA, Witsenburg M, ten Harkel AD. Implantable cardioverter defibrillator therapy for prevention of sudden cardiac death in children in the Netherlands. Pacing Clin Electrophysiol. 2010;33:179–185. doi: 10.1111/j.1540-8159.2009.02603.x

5. Aykan HH, Karagoz T, Gulgun M, Ertugrul I, Aypar E, Ozer S, Alehan D, Celiker A, Ozkutlu S. Midterm Results of Implantable Cardioverter Defibrillators in Children and Young Adults from a Single Center in Turkey. Pacing Clin Electrophysiol. 2016;39:1225–1239. doi: 10.1111/pace.12954

6. Berul CI, Van Hare GF, Kertesz NJ, Dubin AM, Cecchin F, Collins KK, Cannon BC, Alexander ME, Triedman JK, Walsh EP, et al. Results of a multicenter retrospective implantable cardioverter-defibrillator registry of pediatric and congenital heart disease patients. J Am Coll Cardiol. 2008;51:1685–1691. doi: 10.1016/j.jacc.2008.01.033

7. DeWitt ES, Triedman JK, Cecchin F, Mah DY, Abrams DJ, Walsh EP, Gauvreau K, Alexander ME. Time dependence of risks and benefits in pediatric primary prevention implantable cardioverter-defibrillator therapy. Circ Arrhythm Electrophysiol. 2014;7:1057–1063. doi: 10.1161/CIRCEP.114.001569

8. Garnreiter JM, Pilcher TA, Etheridge SP, Saarel EV. Inappropriate ICD shocks in pediatrics and congenital heart disease patients: Risk factors and programming strategies. Heart Rhythm. 2015;12:937–942. doi: 10.1016/j.hrthm.2015.01.028

9. Lawrence D, Von Bergen N, Law IH, Bradley DJ, Dick M, 2nd, Frias PA, Streiper MJ, Fischbach PS. Inappropriate ICD discharges in single-chamber versus dual-chamber devices in the pediatric and young adult population. J Cardiovasc Electrophysiol. 2009;20:287–290. doi: 10.1111/j.1540-8167.2008.01322.x

10. Lewandowski M, Sterlinski M, Maciag A, Syska P, Kowalik I, Szwed H, Chojnowska L, Przybylski A. Long-term follow-up of children and young adults treated with implantable cardioverter-defibrillator: the authors’ own experience with optimal implantable cardioverter-defibrillator programming. Europace. 2010;12:1245–1250. doi: 10.1093/europace/euq263

11. Norrish G, Chubb H, Field E, McLeod K, Ilina M, Spentzou G, Till J, Daubeney PEF, Stuart AG, Matthews J, et al. Clinical outcomes and programming strategies of implantable cardioverter-defibrillator devices in paediatric hypertrophic cardiomyopathy: a UK National Cohort Study. Europace. 2021;23:400–408. doi: 10.1093/europace/euaa307

12. Song MK, Uhm JS, Baek JS, Yoon JK, Na JY, Yu HT, Yang JH, Oh S, Park SW, Song J, et al. Clinical Outcomes of Implantable Cardioverter-Defibrillator in Pediatric Patients - A Korean Multicenter Study. Circ J. 2021;85:1356–1364. doi: 10.1253/circj.CJ-20-0468

13. Einbinder T, Machtei A, Birk E, Schamroth Pravda N, Frenkel G, Amir G, Fogelman R. Low Risk of Inappropriate Shock Among Pediatric Patients With an Implantable Cardioverter Defibrillator: A Single Center Experience. Pediatr Cardiol. 2023. doi: 10.1007/s00246-023-03280-0

14. Robinson JA, LaPage MJ, Atallah J, Webster G, Miyake CY, Ratnasamy C, Ollberding NJ, Mohan S, Von Bergen NH, Johnsrude CL, et al. Outcomes of Pediatric Patients With Defibrillators Following Initial Presentation With Sudden Cardiac Arrest. Circ Arrhythm Electrophysiol. 2021;14:e008517. doi: 10.1161/CIRCEP.120.008517

15. Lewandowski M, Syska P, Kowalik I, Maciag A, Sterlinski M, Atenska-Pawlowska J, Szwed H. Fifteen years’ experience of implantable cardioverter defibrillator in children and young adults: Mortality and complications study. Pediatr Int. 2018;60:923–930. doi: 10.1111/ped.13660

16. Pyngottu A, Werner H, Lehmann P, Balmer C. Health-Related Quality of Life and Psychological Adjustment of Children and Adolescents with Pacemakers and Implantable Cardioverter Defibrillators: A Systematic Review. Pediatr Cardiol. 2019;40:1–16. doi: 10.1007/s00246-018-2038-x

17. Schneider LM, Wong JJ, Adams R, Bates B, Chen S, Ceresnak SR, Danovsky M, Hanisch D, Motonaga KS, Restrepo M, et al. Posttraumatic stress disorder in pediatric patients with implantable cardioverter-defibrillators and their parents. Heart Rhythm. 2022;19:1524–1529. doi: 10.1016/j.hrthm.2022.06.025

18. Rowin EJ, Sridharan A, Madias C, Firely C, Koethe B, Link MS, Maron MS, Maron BJ. Prediction and Prevention of Sudden Death in Young Patients (<20 years) With Hypertrophic Cardiomyopathy. Am J Cardiol. 2020;128:75–83. doi: 10.1016/j.amjcard.2020.04.042

19. Silvetti MS, Tamburri I, Campisi M, Saputo FA, Cazzoli I, Cantarutti N, Cicenia M, Adorisio R, Baban A, Rava L, et al. ICD Outcome in Pediatric Cardiomyopathies. J Cardiovasc Dev Dis. 2022;9. doi: 10.3390/jcdd9020033

20. Cay S, Canpolat U, Ucar F, Ozeke O, Ozcan F, Topaloglu S, Aras D. Programming implantable cardioverter-defibrillator therapy zones to high ranges to prevent delivery of inappropriate device therapies in patients with primary prevention: results from the RISSY-ICD (Reduction of Inappropriate ShockS bY InCreaseD zones) trial. Am J Cardiol. 2015;115:1235–1243. doi: 10.1016/j.amjcard.2015.01.558

21. Moss AJ, Schuger C, Beck CA, Brown MW, Cannom DS, Daubert JP, Estes NA, 3rd, Greenberg H, Hall WJ, Huang DT, et al. Reduction in inappropriate therapy and mortality through ICD programming. N Engl J Med. 2012;367:2275–2283. doi: 10.1056/NEJMoa1211107

22. Ruwald AC, Schuger C, Moss AJ, Kutyifa V, Olshansky B, Greenberg H, Cannom DS, Estes NA, Ruwald MH, Huang DT, et al. Mortality reduction in relation to implantable cardioverter defibrillator programming in the Multicenter Automatic Defibrillator Implantation Trial-Reduce Inappropriate Therapy (MADIT-RIT). Circ Arrhythm Electrophysiol. 2014;7:785–792. doi: 10.1161/CIRCEP.114.001623

23. Guedon-Moreau L, Lacroix D, Sadoul N, Clementy J, Kouakam C, Hermida JS, Aliot E, Boursier M, Bizeau O, Kacet S, et al. A randomized study of remote follow-up of implantable cardioverter defibrillators: safety and efficacy report of the ECOST trial. Eur Heart J. 2013;34:605–614. doi: 10.1093/eurheartj/ehs425

24. Ploux S, Swerdlow CD, Strik M, Welte N, Klotz N, Ritter P, Haissaguerre M, Bordachar P. Towards eradication of inappropriate therapies for ICD lead failure by combining comprehensive remote monitoring and lead noise alerts. J Cardiovasc Electrophysiol. 2018;29:1125–1134. doi: 10.1111/jce.13653

25. Varma N, Epstein AE, Irimpen A, Schweikert R, Love C, Investigators T. Efficacy and safety of automatic remote monitoring for implantable cardioverter-defibrillator follow-up: the Lumos-T Safely Reduces Routine Office Device Follow-up (TRUST) trial. Circulation. 2010;122:325–332. doi: 10.1161/CIRCULATIONAHA.110.937409

26. Boyer SL, Silka MJ, Bar-Cohen Y. Current practices in the monitoring of cardiac rhythm devices in pediatrics and congenital heart disease. Pediatr Cardiol. 2015;36:821–826. doi: 10.1007/s00246-014-1090-4

27. de Asmundis C, Ricciardi D, Namdar M, Chierchia GB, Sarkozy A, Brugada P. Role of home monitoring in children with implantable cardioverter defibrillators for Brugada syndrome. Europace. 2013;15 Suppl 1:i17–i25. doi: 10.1093/europace/eut112

28. Malloy LE, Gingerich J, Olson MD, Atkins DL. Remote monitoring of cardiovascular implantable devices in the pediatric population improves detection of adverse events. Pediatr Cardiol. 2014;35:301–306. doi: 10.1007/s00246-013-0774-5

29. Dechert BE, Bradley DJ, Serwer GA, Dick M, 2nd, LaPage MJ. Frequency of CIED remote monitoring: A quality improvement follow-up study. Pacing Clin Electrophysiol. 2019;42:959–962. doi: 10.1111/pace.13707

30. Tsao S. What is the optimal remote monitoring schedule in pediatric patients with cardiovascular implantable electronic devices? Pacing Clin Electrophysiol. 2019;42:963–964. doi: 10.1111/pace.13706

31. Ananwattanasuk T, Tanawuttiwat T, Chokesuwattanaskul R, Lathkar-Pradhan S, Barham W, Oral H, Thakur RK, Jongnarangsin K. Programming implantable cardioverter-defibrillator in primary prevention: Guideline concordance and outcomes. Heart Rhythm. 2020;17:1101–1106. doi: 10.1016/j.hrthm.2020.02.004

32. Stiles MK, Fauchier L, Morillo CA, Wilkoff BL. 2019 HRS/EHRA/APHRS/LAHRS focused update to 2015 expert consensus statement on optimal implantable cardioverter-defibrillator programming and testing. Heart Rhythm. 2020;17:e220–e228. doi: 10.1016/j.hrthm.2019.02.034

33. Siddiqui O. Statistical methods to analyze adverse events data of randomized clinical trials. J Biopharm Stat. 2009;19:889–899. doi: 10.1080/10543400903105463

34. Nelson W. Confidence-Limits for Recurrence Data - Applied to Cost or Number of Product Repairs. Technometrics. 1995;37:147–157. doi: Doi 10.2307/1269616

35. Olde Nordkamp LR, Postema PG, Knops RE, van Dijk N, Limpens J, Wilde AA, de Groot JR. Implantable cardioverter-defibrillator harm in young patients with inherited arrhythmia syndromes: A systematic review and meta-analysis of inappropriate shocks and complications. Heart Rhythm. 2016;13:443–454. doi: 10.1016/j.hrthm.2015.09.010

36. Gasparini M, Lunati MG, Proclemer A, Arenal A, Kloppe A, Martinez Ferrer JB, Hersi AS, Gulaj M, Wijffels MCE, Santi E, et al. Long Detection Programming in Single-Chamber Defibrillators Reduces Unnecessary Therapies and Mortality: The ADVANCE III Trial. JACC Clin Electrophysiol. 2017;3:1275–1282. doi: 10.1016/j.jacep.2017.05.001

37. Parthiban N, Esterman A, Mahajan R, Twomey DJ, Pathak RK, Lau DH, Roberts-Thomson KC, Young GD, Sanders P, Ganesan AN. Remote Monitoring of Implantable Cardioverter-Defibrillators: A Systematic Review and Meta-Analysis of Clinical Outcomes. J Am Coll Cardiol. 2015;65:2591–2600. doi: 10.1016/j.jacc.2015.04.029

38. Dechert BE, Serwer GA, Bradley DJ, Dick M, 2nd, LaPage MJ. Cardiac implantable electronic device remote monitoring surveillance in pediatric and congenital heart disease: Utility relative to frequency. Heart Rhythm. 2015;12:117–122. doi: 10.1016/j.hrthm.2014.10.009

39. van der Werf C, Lieve KV, Bos JM, Lane CM, Denjoy I, Roses-Noguer F, Aiba T, Wada Y, Ingles J, Leren IS, et al. Implantable cardioverter-defibrillators in previously undiagnosed patients with catecholaminergic polymorphic ventricular tachycardia resuscitated from sudden cardiac arrest. Eur Heart J. 2019;40:2953–2961. doi: 10.1093/eurheartj/ehz309

40. Bergeman AT, Lieve KVV, Kallas D, Bos JM, Roses INF, Denjoy I, Zorio E, Kammeraad JAE, Peltenburg PJ, Tobert K, et al. Flecainide Is Associated With a Lower Incidence of Arrhythmic Events in a Large Cohort of Patients With Catecholaminergic Polymorphic Ventricular Tachycardia. Circulation. 2023;148:2029–2037. doi: 10.1161/CIRCULATIONAHA.123.064786

41. Bergau L, Willems R, Sprenkeler DJ, Fischer TH, Flevari P, Hasenfuss G, Katsaras D, Kirova A, Lehnart SE, Luthje L, et al. Differential multivariable risk prediction of appropriate shock versus competing mortality - A prospective cohort study to estimate benefits from ICD therapy. Int J Cardiol. 2018;272:102–107. doi: 10.1016/j.ijcard.2018.06.103

